# Anti-Mi2 autoantibodies target the PHD fingers of SP140L and TIF1γ, while anti-TIF1γ autoantibodies primarily recognize epitopes outside the PHD region of TIF1γ

**DOI:** 10.1101/2025.03.04.25323364

**Authors:** Iago Pinal-Fernandez, Jon Musai, Maria Casal-Dominguez, Katherine Pak, Mariana Kaplan, Blake M. Warner, Lisa G. Rider, Rohit Aggarwal, Chester V. Oddis, Siamak Moghadam-Kia, Gloria Garrabou, Albert Selva-O’Callaghan, Jose C. Milisenda, John A. Chiorini, Andrew L. Mammen, Peter D. Burbelo

**Affiliations:** Muscle Disease Section, National Institute of Arthritis and Musculoskeletal and Skin Disease, National Institutes of Health, Bethesda, MD, USA; Johns Hopkins University School of Medicine, Baltimore, MD, USA; Systemic Autoimmunity Branch, National Institute of Arthritis and Musculoskeletal and Skin Diseases, National Institutes of Health, Bethesda, MD, USA; Salivary Disorders Unit, National Institute of Dental and Craniofacial Research, National Institutes of Health, Bethesda, MD, USA; Environmental Autoimmunity Group, Clinical Research Branch, National Institute of Environmental Health Sciences, National Institutes of Health, Bethesda, MD, USA; Division of Rheumatology, University of Pittsburgh, Pittsburgh, PA; Muscle Research Unit, Internal Medicine Service, Hospital Clinic, Barcelona, Spain; Barcelona University, Barcelona, Spain; CIBERER and IDIBAPS, Barcelona, Spain; Systemic Autoimmune Disease Unit, Vall d’Hebron Institute of Research, Barcelona, Spain; Autonomous University of Barcelona, Barcelona, Spain; Adeno-Associated Virus Biology Section, National Institute of Dental and Craniofacial Research, National Institutes of Health, Bethesda, MD, USA

**Author notes:** **Address correspondence to**: Dr. Peter D. Burbelo, National Institute of Dental and Craniofacial Research, National Institutes of Health, 10 Center Dr., Bldg. 10, Rm. 1A01, Bethesda, MD 20892. Andrew L. Mammen M.D., Ph.D., or Iago Pinal-Fernandez M.D., Ph.D., Ph.D., Muscle Disease Section, National Institute of Arthritis and Musculoskeletal and Skin Diseases, National Institutes of Health, 50 South Drive, Room 1141, Building 50, MSC 8024, Bethesda, MD 20892. These authors contributed equally to this project (Iago Pinal-Fernandez, Jon Musai).

**Keywords:** Myositis, dermatomyositis, autoantibodies, anti-Mi2, anti-TIF1 γ, PHD, zinc finger domain

## Abstract

**Objectives:** Plant homeodomain (PHD) fingers are present in many chromatin-binding proteins. We recently discovered that anti-Mi2 autoantibodies recognize PHD fingers in Mi2 and AIRE. The purpose of this study was to characterize anti-Mi2 autoantibody recognition of PHD fingers in SP140L and TIF1γ as well as to explore recognition of TIF1γ by both anti-TIF1γ and anti-Mi2 autoantibodies.

**Methods:** Luciferase immunoprecipitation system (LIPS) assays were performed to detect autoantibodies against full-length and protein fragments of SP140L and TIF1γ in serum samples from myositis patients, disease controls, and healthy controls.

**Results:** Anti-Mi2 autoantibodies recognized SP140L. When a 49 amino acid fragment of the PHD finger of SP140L was used as the target, the specificity for selectively detecting anti-Mi2 autoantibodies increased. Additionally, anti-Mi2 autoantibodies weakly bound TIF1γ compared to anti-TIF1γ autoantibodies. Excluding the TIF1γ PHD finger from the TIF1γ target autoantigen eliminated cross-reactivity with anti-Mi2 autoantibodies, confirming that anti-Mi2 autoantibodies specifically target the PHD finger of TIF1γ. Switching two amino acids in the TIF1γ PHD finger to resemble those in AIRE markedly enhanced anti-Mi2 autoantibody immunoreactivity. Anti-TIF1γ autoantibodies primarily recognized the N-terminal fragment outside of the PHD finger, indicating this region contains the immunodominant epitopes.

**Conclusions:** Anti-Mi2 autoantibodies recognize the PHD fingers of SP140L and TIF1γ. TIF1γ is recognized by two different myositis-specific autoantibodies: anti-Mi2 autoantibodies bind the C-terminal PHD domain and anti-TIF1γ autoantibodies predominantly bind the N-terminal region. Removing the PHD finger from the anti-TIF1γ target autoantigen can improve the specificity of anti-TIF1γ autoantibody assays by reducing cross-reactivity with anti-Mi2 autoantibodies.

## INTRODUCTION

Myositis is a heterogenous family of autoimmune diseases that includes dermatomyositis (DM), the antisynthetase syndrome (ASyS), immune-mediated necrotizing myopathy (IMNM), and inclusion body myositis (IBM).(1, 2) Most DM patients have a single myositis-specific autoantibody (MSA) targeting an intracellular autoantigen. The most prevalent MSAs in DM are anti-Mi2,(3) anti-NXP2,(4) anti-TIF1Δ,(5) and anti-MDA5.(6) Importantly, each MSA is associated with a unique disease phenotype. For instance, adult patients with anti-TIF1Δ autoantibodies exhibit a significantly heightened risk of malignancy compared to other DM patients but have a low propensity for developing interstitial lung disease (ILD).(7) Conversely, those with MDA5 autoantibodies frequently have ILD but only rarely have a coexisting cancer.(3) While each DM patient is thought to have only a single MSA, anti-Mi2 autoantibodies have shown low-level binding to TIF1γ, posing challenges for the development of a specific immunologic assay for anti-TIF1γ autoantibodies.(8)

We recently demonstrated that anti-Mi2 autoantibodies contribute to DM disease pathogenesis by accumulating in the nuclei of muscle cells where they disrupt the normal ability of the Mi2-containing nucleosome remodeling and deacetylation (NuRD) complex to repress gene expression. As a consequence, muscle tissue from anti-Mi2-positive DM patients has a unique transcriptomic profile characterized by the aberrant expression of genes not normally expressed in muscle cells.(9–11) We subsequently showed that anti-Mi2 autoantibodies bind to the plant homeodomain (PHD) finger of Mi2.(12) These 50-80 amino acid regions are found in many chromatin-binding proteins and mediate their interaction with histones, thereby allowing them to regulate gene expression.(13, 14) Taken together, these observations suggest that anti-Mi2 autoantibodies cause the derepression of gene expression in DM muscle by binding to the PHD finger of Mi2 and interfering with the ability of the Mi2/NuRD complex to bind chromatin.

PHD fingers are found in more than 70 proteins, including autoimmune regulator (AIRE) protein(15), which we previously demonstrated is a target of anti-Mi2 autoantibodies.(12) Intriguingly, the TIF1γ protein also includes a PHD finger, suggesting that anti-Mi2 autoantibodies might cross-react with TIF1γ due to their recognition of this domain.

In this study, we aimed to explore the breadth of anti-Mi2 immunoreactivity against other PHD finger-containing proteins, including TIF1γ and SP140L. We also sought to clarify the dual targeting of TIF1γ by both anti-TIF1γ and anti-Mi2 autoantibodies.

## METHODS

### Patients and Serum Samples

Two different cohorts were used to study autoantibodies. The first cohort, comprising 184 patients and controls (myositis, disease controls, and healthy controls), was designed to test and validate whether the PHD domain of SP140L is specifically recognized by anti-Mi2 autoantibodies. The second cohort, including 134 samples from healthy controls, anti-Mi2, and anti-TIF1γ patients, was used to investigate the immunodominant epitopes of anti-Mi2 and anti-TIF1γ in the TIF1γ protein (Supplementary Table 1). The healthy controls and anti-Mi2 patients included in both cohorts were the same.

Myositis serum samples were obtained from patients who either fulfilled Llyod’s criteria for inclusion body myositis (IBM) (16) or the Casal and Pinal criteria for other types of autoantibody-positive myositis (1). Patients were classified as autoantibody-positive if they tested positive for autoantibodies against Mi2, TIF1γ, NXP2, MDA5, Jo1, HMGCR, or SRP using at least one of the following immunological methods: LIPS, ELISA, *in vitro* transcription and translation followed by immunoprecipitation, line blotting (EUROLINE Autoimmune Inflammatory Myopathies 16 Ag (IgG) test kit), or immunoprecipitation from ^35^S-methionine-labeled cell lysates.

### Standard Protocol Approvals and Patient Consents

Serum samples were collected from patients enrolled in institutional review board (IRB)-approved cohorts at the National Institutes of Health (NIH) in Bethesda, MD, the University of Pittsburgh, PA, and the Clinic and Vall d’Hebron Hospitals in Barcelona, Spain.

The respective studies received IRB approval from the NIH, the University of Pittsburgh, and the Clinic and Vall d’Hebron Hospitals. Written informed consent was obtained from all participants prior to sample collection and study enrollment.

### Anti-AIRE and anti-Mi2**α** LIPS

Luciferase immunoprecipitation system (LIPS) immunoassays were conducted as previously described. cDNAs encoding proteins and protein fragments of interest were cloned in the luciferase expression vector pREN2. The cDNAs for Sp140L and TIF-1g used for generating these plasmids were derived from synthetic DNA obtained from Twist Biosciences (Supplementary Table 2) and included *BamH1* and *Xho1* restriction sites for directional cloning into the pREN2 eukaryotic expression vector as C-terminal *Renilla* luciferase fusion proteins. All constructs were sequenced to confirm their integrity.

The plasmid expression vectors were then transfected into Cos1 cells using Lipofectamine 3000, and crude lysates were harvested 24–48 hours post-transfection. Following centrifugation of the crude cell lysate, the luciferase activity of the clarified supernatant was determined and then employed as target antigens in immunoprecipitation assays. For these autoantibody measurements, serum samples from myositis patients, healthy controls, and individuals with other autoimmune diseases were diluted in assay buffer A (20 mM Tris, pH 7.5; 150 mM NaCl; 5 mM MgCl2; 1% Triton X-100) and 1ml equivalent of each serum sample was incubated in a 96-well microtiter plate for 1 hour along with 10 light units (LU) per well of lysate corresponding to the different luciferase-antigens.. The serum-antigen mixtures were subsequently transferred to microtiter filter plates (Millipore) containing Ultralink protein A/G beads (Invitrogen) and incubated for 1 hour to capture immune complexes. Lastly, the filter plates (Millipore-Sigma) were washed eight times with buffer A and twice with 1× PBS to remove unbound antigens. After the final wash, coelenterazine substrate (Promega) was added to quantify immunoprecipitated Renilla luciferase fusion proteins by measuring light emission (LU) using a Berthold LB 960 Centro microplate luminometer (Berthold Technologies). The cut-off for each construct was defined as the mean value plus five standard deviations of healthy control sera.

### Statistical analysis

Dichotomous variables were summarized as percentages and absolute frequencies. Pairwise comparisons of categorical variables between groups were performed using Fisher’s exact test. For continuous variables, intergroup comparisons were conducted using Student’s t-test.

Sequence homology between protein sequences was analyzed using MUSCLE v3.8 (Multiple Sequence Comparison by Log-Expectation).

All statistical analyses were performed in the R programming language. A two-sided p-value of ≤0.05 was considered statistically significant, with no adjustments made for multiple comparisons.

## RESULTS

### Anti-Mi2 Autoantibodies Recognize the PHD Domain of SP140L

To investigate whether anti-Mi2 autoantibodies also target the PHD fingers of proteins other than Mi2 and AIRE, we identified genes encoding regions of sequence similarity to the PHD finger of Mi2 (https://prosite.expasy.org/PS01359). This analysis identified SP140L and SP140 as potential candidates (Figure 1). Due to the nearly identical protein sequence of this region in SP140L and SP140, we focused on SP140L for further investigation using the LIPS assay.

**Figure 1.**
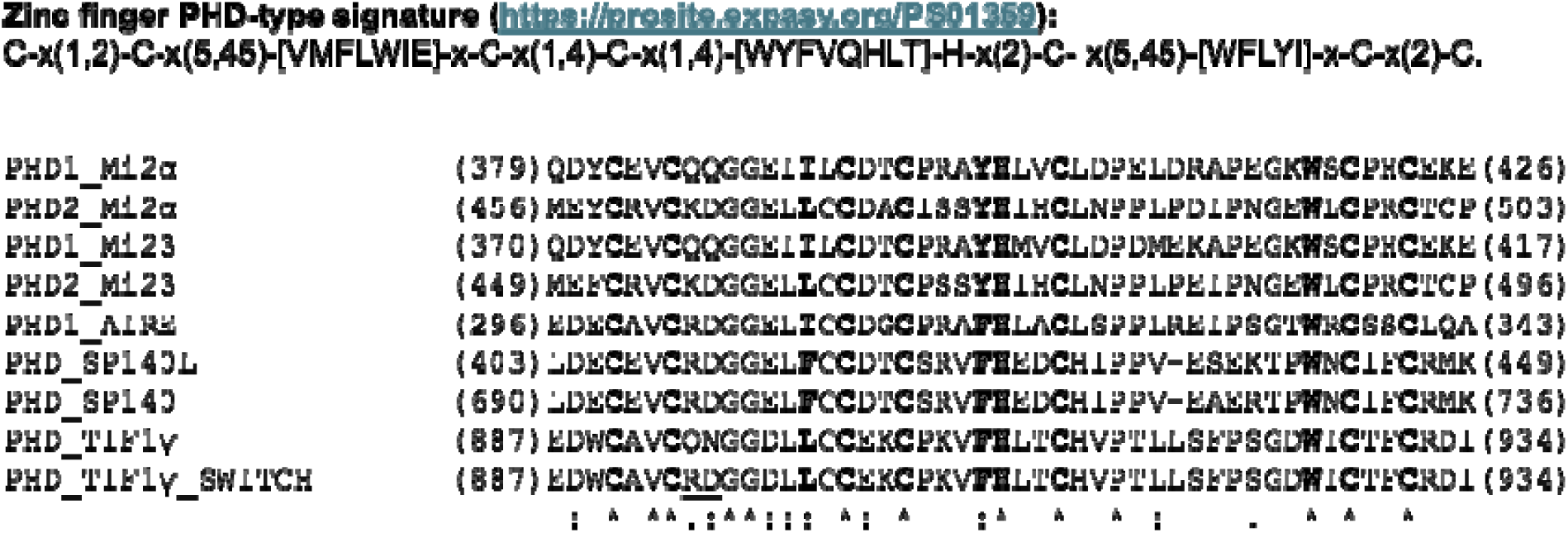
Sequence alignment of the PHD finger domains from Mi2, AIRE, SP140L, SP140, and TIF1γ. The defining motif of the PHD region is highlighted in bold, as detailed below. Starting and ending amino acids are noted at the beginning and end of each sequence. In the modified TIF1γ PHD domain (PHD_TIF1γ_SWITCH), the two amino acids altered to resemble the AIRE and SP140L PHD domains are underlined.

The LIPS assay was used to test this hypothesis with serum samples from healthy controls, myositis patients, and disease controls. As shown in Figure 2a, the full-length SP140L protein in the LIPS assay was predominantly recognized by sera from anti-Mi2 patients across the studied groups. Nevertheless, a small subset of sera from individuals in the other groups also showed recognition of SP140L at levels comparable to those observed in anti-Mi2 patients.

**Figure 2.**
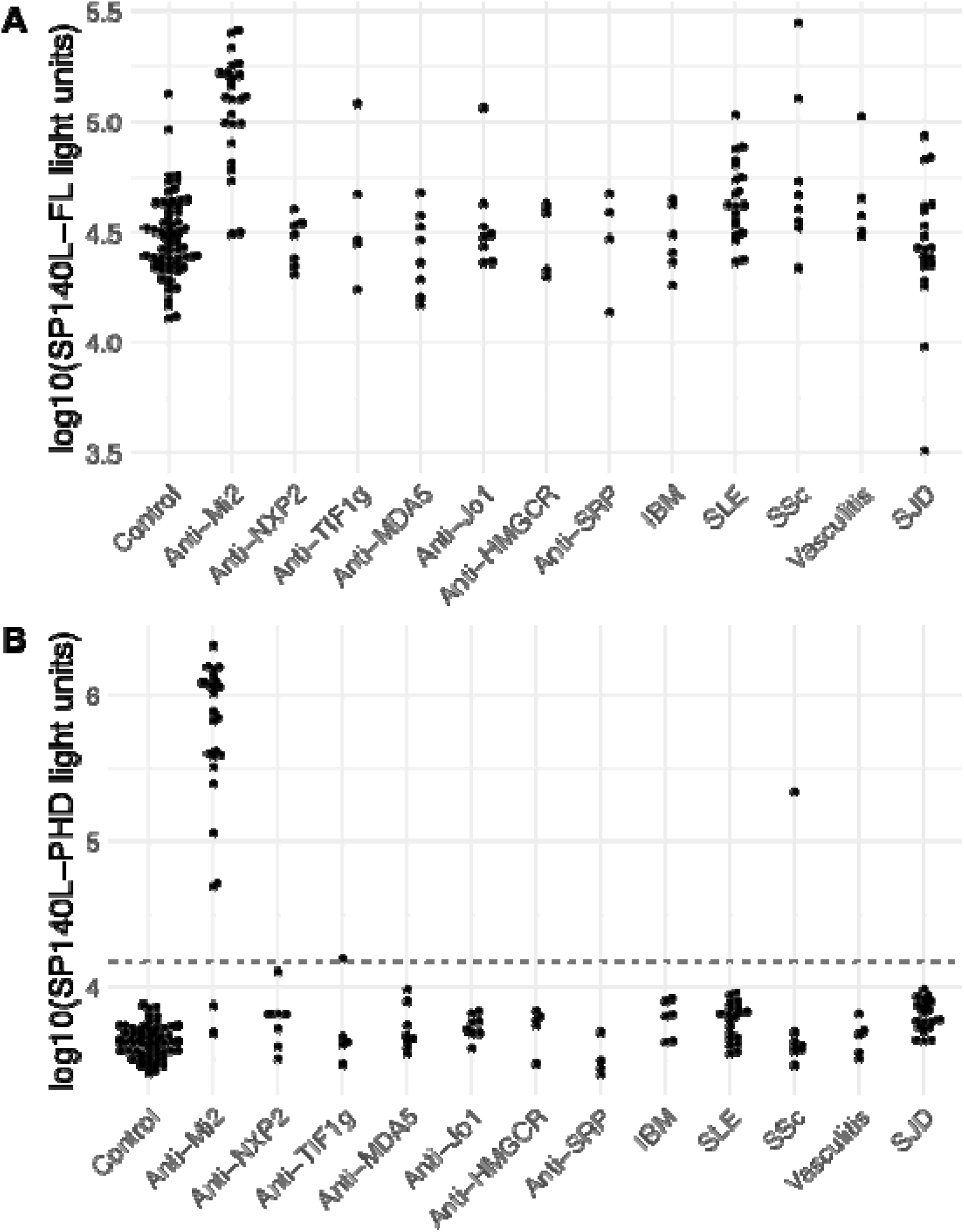
Anti-Mi2 Autoantibodies Recognize the PHD Domain of SP140L. Detection of autoantibodies against the full-length SP140L protein (panel A) and the PHD zinc finger domain of SP140L (panel B) in healthy controls, myositis patients, and individuals with other systemic autoimmune diseases, using the luciferase immunoprecipitation system (LIPS). The dotted line represents the cutoff for anti-AIRE autoantibody positivity, defined as 5 standard deviations above the mean. IBM: inclusion body myositis; SLE: systemic lupus erythematosus; SSc: systemic sclerosis; SJD: Sjögren disease.

To determine the contribution of only the PHD zinc finger domain of SP140L to immunoreactivity, we focused on the conserved 49-amino-acid segment of SP140L (Figure 2b). Testing of this small protein fragment demonstrated all but two anti-Mi2 patient sera (92% sensitivity) were recognized, which exceeded the prespecified cutoff (p=2.2e-16). In contrast to the results seen with the full-length SP140L target antigen, only one anti-TIF1γ sample and one systemic sclerosis sample showed immunoreactivity, resulting in an overall specificity of 99%. These findings highlight that anti-Mi2 autoantibodies specifically recognize the PHD zinc finger domain of SP140L.

### Anti-Mi2 Autoantibodies Recognize the PHD Domain of TIF1**γ**

Based on bioinformatic analysis showing that TIF1γ also contains a related PHD zinc finger domain (Figure 1), we studied immunoreactivity against TIF1γ. Using full-length TIF1γ protein, anti-Mi2 sera showed significant immunoreactivity with TIF1γ compared to sera from healthy controls but showed lower levels than with anti-TIF1γ sera (Figure 3a). As shown in Figure 3a, using the full-length TIF1γ protein to distinguish between anti-TIF1γ and anti-Mi2 autoantibodies presents challenges, necessitating a tradeoff between false-negative anti-TIF1γ results and false-positive anti-Mi2 cases.

**Figure 3.**
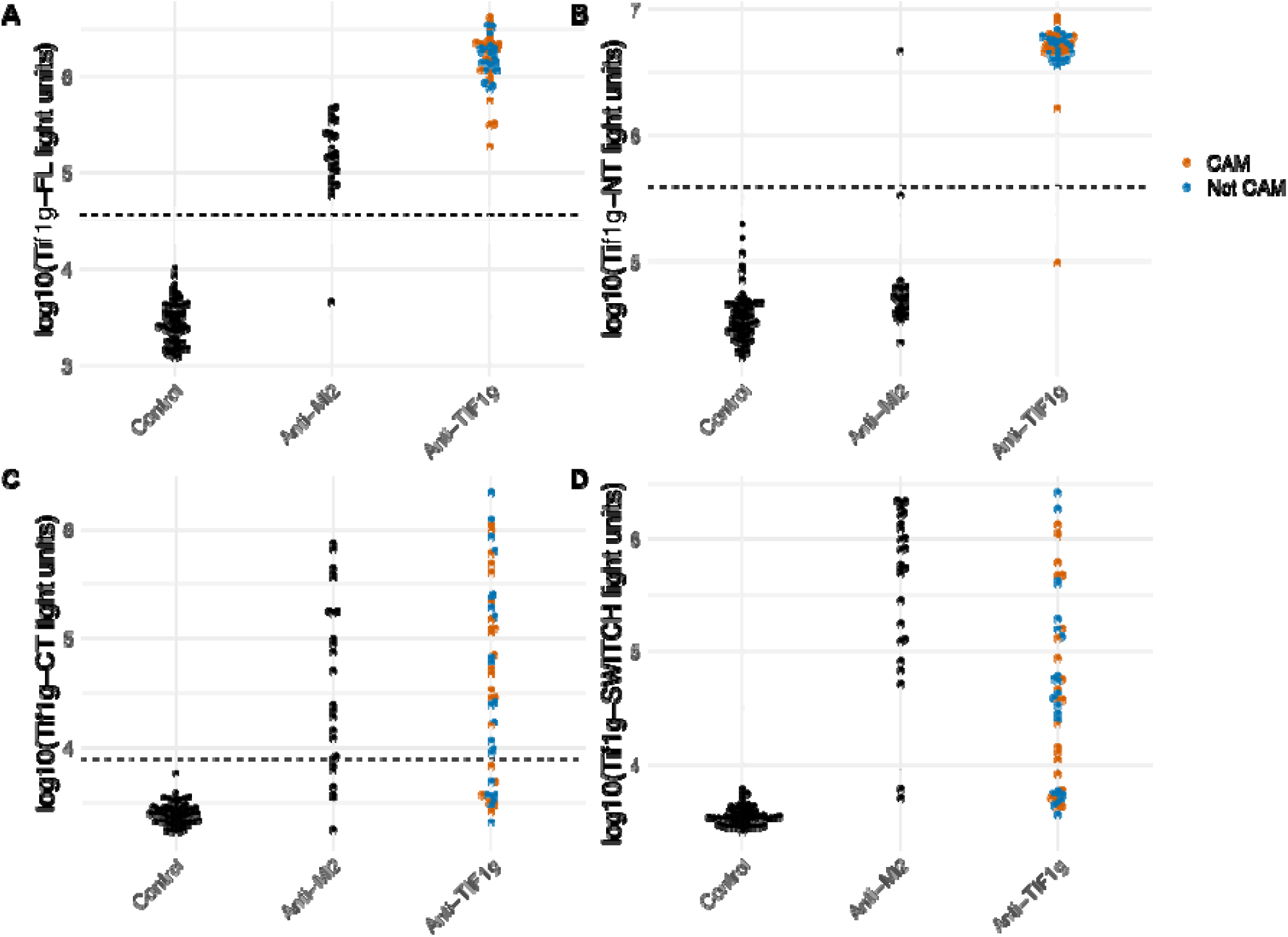
Anti-Mi2 Autoantibodies Recognize the PHD Domain of TIF1γ and anti-TIF1γ Autoantibodies Primarily Recognize Regions Outside the PHD Domain. Detection of autoantibodies was performed using the luciferase immunoprecipitation system (LIPS) against (A) the full-length TIF1γ protein, (B) the N-terminal region of TIF1γ, (C) the PHD-containing C-terminal region of TIF1γ, and (D) a modified C-terminal region of TIF1γ with two amino acid substitutions to increase similarity to AIRE. Autoantibody levels were analyzed in healthy controls and myositis patients with anti-Mi2 or anti-TIF1γ autoantibodies. The dotted line represents the cutoff for anti-TIF1γ autoantibody positivity, defined as 5 standard deviations above the mean. CAM: cancer-associated myositis.

Given the preferential recognition of PHD zinc finger domains by anti-Mi2 autoantibodies, we hypothesized that excluding the PHD domain, located in the C-terminal region of TIF1γ, from TIF1γ in the LIPS assay could improve discrimination between anti-TIF1γ and anti-Mi2 autoantibodies. Indeed, a modified assay using amino acids 2-585 from the N-terminus of TIF1γ (Figure 3b) showed that only one anti-TIF1γ serum sample was misclassified as seronegative, and one anti-Mi2 serum sample was misclassified as seropositive for anti-TIF1γ (p=2.2e-16). Notably, most anti-Mi2 samples had antibody levels similar to those of the healthy controls.

Overall, the modified immunoassay achieved a sensitivity of 98% and a specificity of 99% for anti-TIF1γ autoantibodies, significantly improving its ability to exclusively detect TIF1γ seropositive serum samples.

Additional testing of the C-terminus region (amino acids 586-1127) of the TIF1γ protein showed variable results with immunoreactivity with both anti-TIFγ anti-Mi2 samples (Figure 3c).

To better understand the varying levels of immunoreactivity of anti-Mi2 autoantibodies against the PHD domains across different target proteins, we analyzed the amino acid similarity in detail and examined the crystal structure of the conserved PHD domain in the proteins studied. This analysis identified two discordant amino acid residues in TIF1γ (Q894 and N895) compared to AIRE and SP140L which were located at the apex of the PHD loop. To test the significance of these two amino acid changes, we switched these two amino acids (Q894R, N895D) in the TIF1γ C-terminal protein fragment to match the two residues in AIRE, MI2 and SP140L (Figure 1). This modification resulted in a nearly 10-fold increase in the level of immunoreactivity exclusive to anti-Mi2 samples, without affecting reactivity in anti-TIF1γ samples (p=7.4e-13, Figure 3d and Figure 4). Overall, these results highlight how amino acid differences in the PHD region of these antigens explain the relative levels of immunoreactivity and likely affinity seen against these target proteins.

**Figure 4.**
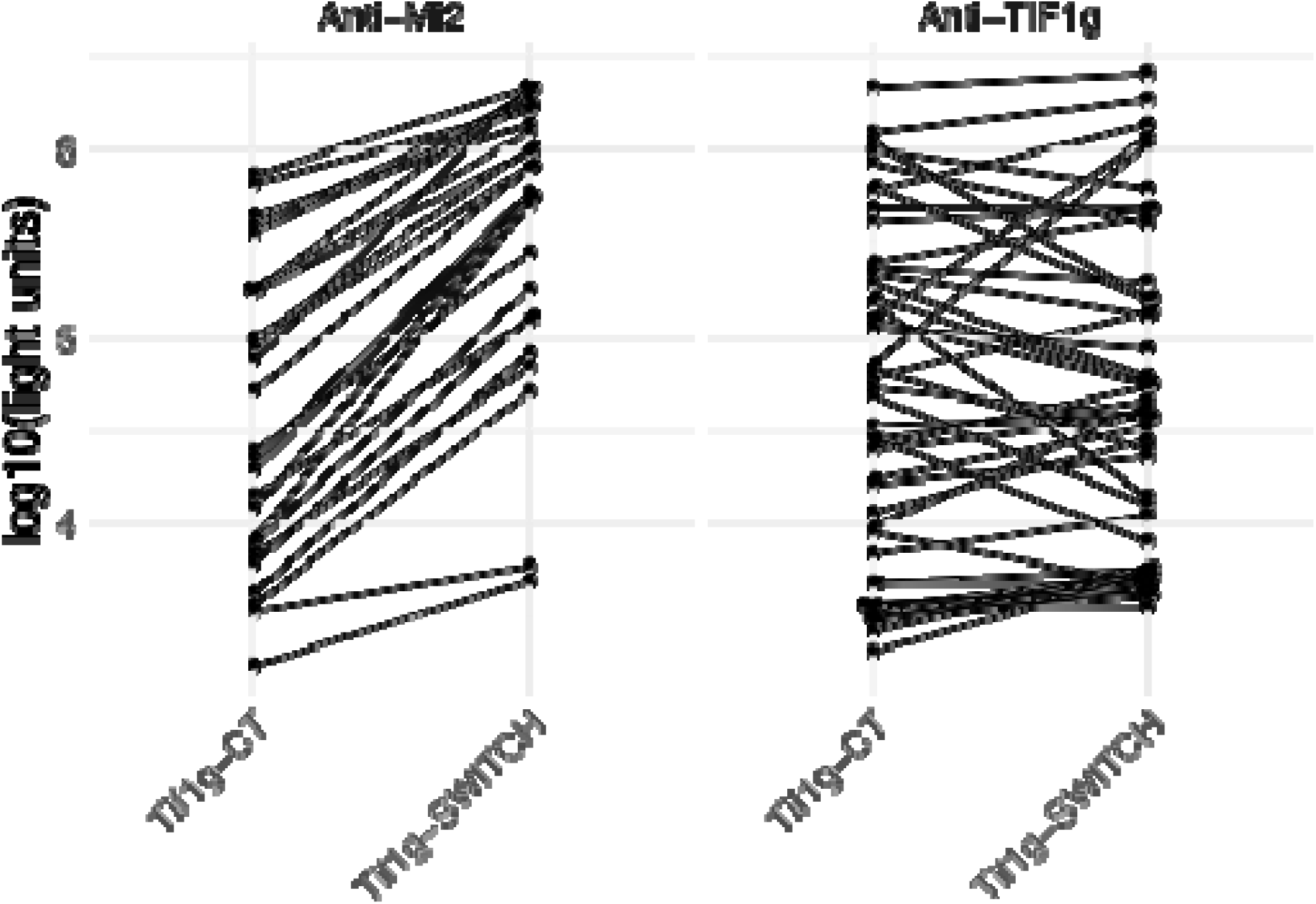
Changing two amino acids in the PHD domain of TIF1γ increases its immunoreactivity for anti-Mi2 autoantibodies but does not alter its recognition by anti-TIF1γ autoantibodies. Autoantibody levels were measured using LIPS against the C-terminal region of TIF1γ (TIF1γ-CT or against the same region containing a 2 amino substitution in the PHD domain (TIF1γ-switch; as described in material and methods) in healthy controls and myositis patients with anti-Mi2 or anti-TIF1γ autoantibodies.

### Anti-TIF1**γ** Autoantibodies Primarily Recognize Regions Outside the PHD Domain, Regardless of Cancer Status

Anti-TIF1γ autoantibodies are strongly associated with cancer and often co-occur with autoantibodies against other proteins such as SP4 and CCAR1 (ref). To further explore immunoreactivity, we used the new assay containing only the N-terminus region of TIF1γ and the results showed a high levels on immunoreactivity regardless of cancer and non-cancer status of the anti-TIF1γ seropositive patients (Figure 3b-c). These findings underscore that anti-TIF1γ autoantibodies predominantly target regions outside the PHD finger, irrespective of cancer status.

## DISCUSSION

In this study, we discovered several key features of anti-Mi2 and anti-TIF1γ autoantibodies. First, we establish that anti-Mi2 autoantibodies bind to the PHD fingers of multiple proteins, including SP140L and TIF1γ. This highlights their broader reactivity profile beyond Mi2, reinforcing the potential to redefine them as primarily “anti-PHD finger” autoantibodies. Second, we show that assays restricted to the PHD finger can significantly improve the diagnostic accuracy for anti-Mi2 autoantibody detection. Third, we identified TIF1γ as a target for multiple myositis-specific autoantibodies, with anti-Mi2 autoantibodies recognizing the PHD finger in the C-terminus of TIF1γ, while anti-TIF1γ autoantibodies primarily bind to the N-terminal region. Finally, we demonstrate that removing the PHD finger from anti-TIF1γ assays enhances specificity by eliminating cross-reactivity with anti-Mi2 autoantibodies.

Anti-Mi2 autoantibodies were first identified by Dr. Reichlin and Dr. Mattioli, who discovered them prior to characterizing their first binding autoantigen, naming them based on the initial patient, “Mi2,” who exhibited this specificity.(21) The first proteins known to be targeted by anti-Mi2 autoantibodies were subsequently named Mi2a and Mi2b. Our findings refine this discovery by showing that anti-Mi2 autoantibodies target other proteins that include PHD fingers, including AIRE, SP140L, and TIF1γ.

The SP140L gene is thought to be a recent evolutionarily member of the SP100 family, formed through DNA rearrangements of the neighboring genes of SP100 and SP140.(22) A small subset of patients with primary biliary cirrhosis have been found to have autoantibodies against SP140L, which is not unexpected given our findings that the full-length protein is recognized by patients with various autoimmune diseases.(22) In contrast, the isolated PHD finger of SP140L demonstrated specific immunoreactivity in patients with anti-Mi2 autoantibodies. Since SP140 and SP140L differ by only two amino acids in their PHD domains, it is likely that SP140 is also a target of anti-Mi2 autoantibodies.

Our prior research has demonstrated that anti-Mi2 autoantibodies can penetrate muscle cells, accumulate in the nucleus, and disrupt the function of their autoantigens.(9, 11) While the Mi2/NuRD complex was the first identified target affected by the internalization of autoantibodies,(9, 11) it is plausible that the dysfunction of other PHD finger-containing proteins also play a role in disease pathogenesis. Investigating these interactions is inherently complex yet fascinating, as it involves unraveling the potential contribution of several PHD finger-containing proteins.

The initial naming of anti-TIF1γ autoantibodies was based on their immunoprecipitation patterns (anti-155/140)(5) and later revised to reflect their target protein, TIF1γ.(23) Our findings challenge the accuracy of this nomenclature, as TIF1γ is targeted not only by anti-TIF1γ autoantibodies but also by anti-Mi2 (i.e., anti-PHD finger) autoantibodies.

Given TIF1γ’s role as a suppressor of IFNβ1 expression in the nucleus, and the central importance of IFNβ1 signaling across all DM autoantibody groups,(11, 24) it is intriguing to speculate that anti-PHD finger autoantibody internalization may block TIF1γ’s inhibitory function, leading to the overexpression of interferon beta. Anti-TIF1γ autoantibodies preferentially target the N-terminal region of TIF1γ, which contains RING and B-box zinc fingers, while anti-Mi2 autoantibodies target the PHD finger. Our study opens the possibility that the overexpression of IFNβ1 in both anti-Mi2-positive and anti-TIF1γ-positive DM may result from the inhibition of TIF1γ function through distinct mechanisms: interaction with the RING and/or B-box zinc finger domains in anti-TIF1γ-positive DM and with the PHD finger in anti-Mi2-positive DM.

Practically, our findings provide a clear path for improving diagnostic accuracy in anti-Mi2 and anti-TIF1γ autoantibody testing. For anti-Mi2 autoantibodies, assays can be optimized by focusing on the PHD finger, while for anti-TIF1γ autoantibodies, excluding the PHD finger from test antigens can reduce cross-reactivity. These strategies can enhance the diagnostic performance of the assays, facilitating the accurate differentiation of these autoantibodies in clinical settings.

Despite these advances, several key questions remain unresolved. For instance, a comprehensive understanding of potential differences in anti-TIF1γ autoantibody binding between cancer and non-cancer patients, particularly within the N-terminal region of TIF1γ, is still lacking. Additionally, although our focus has been on a subset of PHD finger-containing proteins—Mi2, AIRE, SP140L, and TIF1γ—many others remain unexplored. Finally, the functional implications of these findings, including the molecular mechanisms underlying disease in DM patients with anti-TIF1γ autoantibodies and the relationship between autoantibody binding sites and clinical manifestations, require further investigation.

In conclusion, this study enhances our understanding of anti-Mi2 autoantibodies as anti-PHD finger autoantibodies, clarifies the differential recognition of TIF1γ by anti-Mi2 and anti-TIF1γ autoantibodies, and provides strategies to improve the diagnostic accuracy for both. These findings advance our knowledge of the molecular mechanisms underlying myositis and pave the way for more precise diagnostic and therapeutic approaches.

## Contributorship

All authors contributed to the development of the manuscript, including interpretation of results, substantive review of drafts, and approval of the final draft for submission.

## Data Availability

All data produced in the present study are available upon reasonable request to the authors

## Acknowledgments

We thank Ira Targoff for performance of myositis autoantibody testing.

## Ethical approval information

All biospecimens were from subjects enrolled in institutional review board (IRB)-approved cohorts in the National Institutes of Health, the Clinic Hospital, the Vall d’Hebron Hospital.

## Data sharing statement

Any anonymized data not published within the article will be shared by request from any investigator.

## Declaration of generative AI and AI-assisted technologies in the writing process

During the preparation of this work, the author(s) utilized ChatGPT to enhance sentence clarity. Following its use, the author(s) thoroughly reviewed and edited the content as necessary, taking full responsibility for the final version of the publication.

## REFERENCES

1. Casal-Dominguez M, Pinal-Fernandez I, Pak K, Huang W, Selva-O’Callaghan A, Albayda J, et al. Performance of the 2017 European Alliance of Associations for Rheumatology/American College of Rheumatology Classification Criteria for Idiopathic Inflammatory Myopathies in Patients With Myositis-Specific Autoantibodies. Arthritis Rheumatol. 2022;74(3):508-17.

2. Selva-O’Callaghan A, Pinal-Fernandez I, Trallero-Araguas E, Milisenda JC, Grau-Junyent JM, Mammen AL. Classification and management of adult inflammatory myopathies. Lancet Neurol. 2018;17(9):816–28.

3. Pinal-Fernandez I, Mecoli CA, Casal-Dominguez M, Pak K, Hosono Y, Huapaya J, et al. More prominent muscle involvement in patients with dermatomyositis with anti-Mi2 autoantibodies. Neurology. 2019;93(19):e1768–e77.

4. Albayda J, Pinal-Fernandez I, Huang W, Parks C, Paik J, Casciola-Rosen L, et al. Antinuclear Matrix Protein 2 Autoantibodies and Edema, Muscle Disease, and Malignancy Risk in Dermatomyositis Patients. Arthritis Care Res (Hoboken). 2017;69(11):1771–6.

5. Targoff IN, Mamyrova G, Trieu EP, Perurena O, Koneru B, O’Hanlon TP, et al. A novel autoantibody to a 155-kd protein is associated with dermatomyositis. Arthritis Rheum. 2006;54(11):3682–9.

6. Sato S, Hoshino K, Satoh T, Fujita T, Kawakami Y, Fujita T, et al. RNA helicase encoded by melanoma differentiation-associated gene 5 is a major autoantigen in patients with clinically amyopathic dermatomyositis: Association with rapidly progressive interstitial lung disease. Arthritis Rheum. 2009;60(7):2193–200.

7. Trallero-Araguas E, Rodrigo-Pendas JA, Selva-O’Callaghan A, Martinez-Gomez X, Bosch X, Labrador-Horrillo M, et al. Usefulness of anti-p155 autoantibody for diagnosing cancer-associated dermatomyositis: a systematic review and meta-analysis. Arthritis Rheum. 2012;64(2):523–32.

8. Fujimoto M, Murakami A, Kurei S, Okiyama N, Kawakami A, Mishima M, et al. Enzyme-linked immunosorbent assays for detection of anti-transcriptional intermediary factor-1 gamma and anti-Mi-2 autoantibodies in dermatomyositis. J Dermatol Sci. 2016;84(3):272–81.

9. Pinal-Fernandez I, Milisenda JC, Pak K, Munoz-Braceras S, Casal-Dominguez M, Torres-Ruiz J, et al. Transcriptional derepression of CHD4/NuRD-regulated genes in the muscle of patients with dermatomyositis and anti-Mi2 autoantibodies. Ann Rheum Dis. 2023;82(8):1091–7.

10. Pinal-Fernandez I, Casal-Dominguez M, Derfoul A, Pak K, Miller FW, Milisenda JC, et al. Machine learning algorithms reveal unique gene expression profiles in muscle biopsies from patients with different types of myositis. Ann Rheum Dis. 2020;79(9):1234–42.

11. Pinal-Fernandez I, Munoz-Braceras S, Casal-Dominguez M, Pak K, Torres-Ruiz J, Musai J, et al. Pathological autoantibody internalisation in myositis. Ann Rheum Dis. 2024;83(11):1549–60.

12. Musai J, Jayaraman S, Pak K, Pinal-Fernandez I, Munoz-Braceras S, Casal-Dominguez M, et al. Myositis-specific autoantibodies recognizing Mi2 also target the autoimmune regulator (AIRE) protein at a shared PHD-zinc finger. bioRxiv. 2025.

13. Koh AS, Kuo AJ, Park SY, Cheung P, Abramson J, Bua D, et al. Aire employs a histone-binding module to mediate immunological tolerance, linking chromatin regulation with organ-specific autoimmunity. Proc Natl Acad Sci U S A. 2008;105(41):15878–83.

14. Musselman CA, Ramirez J, Sims JK, Mansfield RE, Oliver SS, Denu JM, et al. Bivalent recognition of nucleosomes by the tandem PHD fingers of the CHD4 ATPase is required for CHD4-mediated repression. Proc Natl Acad Sci U S A. 2012;109(3):787–92.

15. Jain K, Fraser CS, Marunde MR, Parker MM, Sagum C, Burg JM, et al. Characterization of the plant homeodomain (PHD) reader family for their histone tail interactions. Epigenetics Chromatin. 2020;13(1):3.

16. Lloyd TE, Mammen AL, Amato AA, Weiss MD, Needham M, Greenberg SA. Evaluation and construction of diagnostic criteria for inclusion body myositis. Neurology. 2014;83(5):426–33.

17. Burbelo PD, Ching KH, Klimavicz CM, Iadarola MJ. Antibody profiling by Luciferase Immunoprecipitation Systems (LIPS). J Vis Exp. 2009(32).

18. Burbelo PD, Riedo FX, Morishima C, Rawlings S, Smith D, Das S, et al. Sensitivity in Detection of Antibodies to Nucleocapsid and Spike Proteins of Severe Acute Respiratory Syndrome Coronavirus 2 in Patients With Coronavirus Disease 2019. J Infect Dis. 2020;222(2):206–13.

19. Burbelo PD, Iadarola MJ, Keller JM, Warner BM. Autoantibodies Targeting Intracellular and Extracellular Proteins in Autoimmunity. Front Immunol. 2021;12:548469.

20. Burbelo PD, Huapaya JA, Khavandgar Z, Beach M, Pinal-Fernandez I, Mammen AL, et al. Quantification of autoantibodies using a luminescent profiling method in autoimmune interstitial lung disease. Front Immunol. 2024;15:1462242.

21. Reichlin M, Mattioli M. Description of a serological reaction characteristic of polymyositis. Clin Immunol Immunopathol. 1976;5(1):12–20.

22. Saare M, Hamarik U, Venta R, Panarina M, Zucchelli C, Pihlap M, et al. SP140L, an Evolutionarily Recent Member of the SP100 Family, Is an Autoantigen in Primary Biliary Cirrhosis. J Immunol Res. 2015;2015:526518.

23. Fujimoto M, Hamaguchi Y, Kaji K, Matsushita T, Ichimura Y, Kodera M, et al. Myositis-specific anti-155/140 autoantibodies target transcription intermediary factor 1 family proteins. Arthritis Rheum. 2012;64(2):513–22.

24. Pinal-Fernandez I, Casal-Dominguez M, Derfoul A, Pak K, Plotz P, Miller FW, et al. Identification of distinctive interferon gene signatures in different types of myositis. Neurology. 2019;93(12):e1193–e204.

